# Defining the Genetic and Phenotypic Landscape of Primary Immune Regulatory Disorders Using the ClinGen Validation Framework

**DOI:** 10.64898/2026.09.11.26362285

**Authors:** Benjamin D. Solomon, Justyne E. Ross, Eleanor P. Fensterle, Rasha S. Soliman, Michelle K. Paczosa, Alison Brittain, Elizabeth M. Forbes, Olga F. Sarmento, Ivana Stojkic, Shifaa Alkotob, Jahnavi Aluri, Jorge Diogo Da Silva, Ana Rita Soares, Monica Sulit, Anita Chandra, Fabian Hauck, Stephen Jolles, Paul J. Maglione, Harry Lesmana, Craig D. Platt, Markus G. Seidel, Andrew L. Snow, Kathleen E. Sullivan, Troy R. Torgerson, Tiphanie P. Vogel, Klaus Warnatz, Kejian Zhang, Purvesh Khatri, Forum Raval, Stuart G. Tangye, Roshini S. Abraham

## Abstract

**Background:** Primary immune regulatory disorders (PIRDs) represent a rapidly growing group of inborn errors of immunity (IEIs) characterized by infection susceptibility, autoimmunity, inflammation, and lymphoproliferation. However, the core features of the gene-disease relationships underlying these conditions are often obscured by limitations in available published data. The Clinical Genome Resource (ClinGen) is an international collaborative effort that seeks to address this challenge for PIRDs and other monogenic diseases through a standardized framework for classifying the strength of gene-disease relationships.

**Objective:** We sought to systematically evaluate the evidence for proposed PIRD gene-disease relationships and identify conserved phenotypic features across this heterogeneous group of disorders.

**Methods:** Using the standardized ClinGen framework, the ClinGen PIRD Gene Curation Expert Panel (GCEP) identified potential gene-disease relationships for monogenic conditions characterized predominantly by immune dysregulation. We subsequently curated evidence relevant to these relationships, classified the strength of evidence for these gene-disease relationships, and evaluated clinical patterns among these conditions through standardized phenotyping using the Human Phenotype Ontology (HPO).

**Results:** As of April 2026, the PIRD-GCEP has curated a total of 46 genes corresponding to 49 gene-disease relationships characterized by immune dysregulation. Of these, 28 were categorized as definitive, 2 as strong, 9 as moderate, 7 as limited, and 3 as disputed. Analysis of HPO-based phenotyping revealed 3 major phenotypic clusters corresponding to lymphoproliferation and systemic inflammation, atopic and gastrointestinal inflammation, and combined immune deficiency with autoimmunity.

**Conclusion:** The PIRD-GCEP framework provides validated gene-disease classifications and identifies three distinct phenotypic clusters, facilitating improved diagnosis while revealing genes requiring further investigation to confirm their role in immune regulatory disorders.

**CAPSULE SUMMARY:** Using the ClinGen framework, this study describes the first global initiative to validate gene-disease relationships for primary immune regulatory disorders, identifying 49 relationships and three phenotypic clusters, providing a resource for genetic testing and diagnosis.

**KEY MESSAGES:**

- The PIRD-GCEP applied ClinGen’s standardized framework to curate 46 genes (49 gene-disease relationships) linked to primary immune regulatory disorders. Thirty of these relationships were classified as definitive or strong, giving clinicians a validated basis for variant interpretation in this heterogeneous disease group.
- Standardized HPO phenotyping revealed that, despite their genetic diversity, PIRDs converge onto a small number of clinically meaningful patterns including: (1) lymphoproliferation with systemic inflammation, (2) atopic and gastrointestinal inflammation, and (3) immune deficiency with autoimmunity.
- This validated set of gene-disease relationships and phenotypic clusters represents a resource for the genetic testing and diagnosis of PIRDs, potentially reducing diagnostic delays and aiding in the recognition of novel PIRD gene-disease relationships.

## INTRODUCTION

Since the International Union of Immunological Societies (IUIS) first began systematically classifying inborn errors of immunity (IEIs) in 1999 ^1^, the number of recognized conditions has grown over five-fold. The most recent 2024 IUIS classification now includes 559 distinct conditions with “diseases of immune dysregulation” representing one of the most rapidly expanding groups ^2,3^. Early descriptions of primary immune disorders primarily emphasized susceptibility to severe and recurrent infections. However, expanding clinical and experimental evidence subsequently demonstrated that monogenic immune diseases also contribute to pathologic inflammation, autoimmunity, lymphoproliferation, and atopy. This broader understanding led to the inclusion of immune dysregulation disorders as a distinct category in the 2004 IUIS update ^4^. Indeed, “improving recognition of immune dysregulation diseases, including the growing field of autoinflammatory disorders and interferonopathies” prompted the transition from the term "primary immune deficiency" to the more comprehensive designation "inborn errors of immunity" in the 2017 IUIS update ^5^.

Primary immune regulatory disorders (PIRDs) comprise a heterogeneous group of conditions characterized by excessive immune activation and impaired resolution of normal inflammatory responses. For example, in autoimmune lymphoproliferative syndrome (ALPS), defective *FAS*-mediated apoptosis results in excessive lymphoproliferation, presenting as lymphadenopathy, splenomegaly, and an increased lifetime risk of lymphoma. Failure of apoptosis also permits self-reactive lymphocytes to evade activation-induced cell death ^6^ and skews B-cell selection ^7,8^, leading to antibody-mediated cytopenias and other autoimmune manifestations. Familial hemophagocytic lymphohistiocytosis (fHLH) represents another well-described group of PIRDs where defects in genes including *PRF1*, *STX11*, *STXBP2*, and *UNC13D* impair the cytotoxic function of NK cells and CD8⁺ T cells ^9^. This defect leads to inadequate viral clearance, persistent CD8+ T cell activation, and sustained IFNγ-driven inflammation that contributes to multi-organ failure. Moreover, impaired cytotoxicity contributes to increased viral susceptibility and malignancy due to impaired immune surveillance ^9^.

However, the rarity and phenotypic heterogeneity of PIRDs can complicate disease classification, variant interpretation, and consistent diagnostic evaluation across institutions. The Clinical Genome Resource (ClinGen), a National Institutes of Health (NIH)-funded initiative, is uniquely positioned to address this challenge through its efforts to systematically evaluate gene-disease relationships (GDRs) and variant pathogenicity through a standardized, evidence-based framework ^10,11^. ClinGen organizes this work through Gene Curation Expert Panels (GCEPs), which comprise clinicians, scientists, and genetic counselors with domain expertise across academic medical centers, research institutions, diagnostic laboratories, and industry. Curation of inborn errors of immunity (IEI) is further coordinated by the Immunology Clinical Domain Working Group (CDWG) ^12^, which includes panels such as the ClinGen Antibody Deficiencies (AD)-GCEP ^13^. By aggregating and curating clinical and experimental data, ClinGen enables rigorous and consistent characterization of PIRDs and other monogenic diseases, improving diagnostic accuracy and guiding clinical management.

Despite the rapid expansion of recognized PIRD-associated genes, no systematic effort has evaluated the validity of these GDRs or defined conserved phenotypic patterns across disorders. Here, we describe the work of the ClinGen PIRD-GCEP to curate GDRs with associated immune dysregulation according to ClinGen’s standardized framework ^14^. These GDRs include those described in IUIS IEI Table 4 (Diseases of Immune Dysregulation ^3^) and others identified by the GCEP’s content domain experts. As of April 2026, we have curated and classified 46 genes related to PIRDs, corresponding to 49 gene-disease relationships. This includes data from over 340 patients with published genotypic and phenotypic data, resulting in over 2,000 phenotypic annotations using the standardized Human Phenotype Ontology (HPO). These annotations allowed us to identify 3 phenotypic clusters broadly corresponding to lymphoproliferation, autoimmunity, and immune deficiency. Together, these data provide a comprehensive landscape of PIRD genotypes and phenotypes.

## METHODS

### ClinGen framework

ClinGen provides standardized operating procedures (SOPs) to systematically evaluate the evidence for gene-disease, variant-pathogenicity, gene dosage, and diagnosis-clinical actionability relationships, as well as an online repository for open dissemination of the resulting data (**Figure 1**). For the gene-disease relationships curated here, the SOP is broadly divided into pre-curation and curation described below. These curation tasks are organized into GCEPs focused on discrete clinical domains and composed of individuals with content expertise in those domains. GCEPs are responsible for determining those genes within the scope of the GCEP’s clinical domain, carrying out the curations, and adjudicating final gene-disease relationship classifications. ClinGen curations are updated periodically; to find the most current information please visit clinicalgenome.org. Information for the PIRD-GCEP can be found at https://clinicalgenome.org/affiliation/40145/.

**Figure 1:**
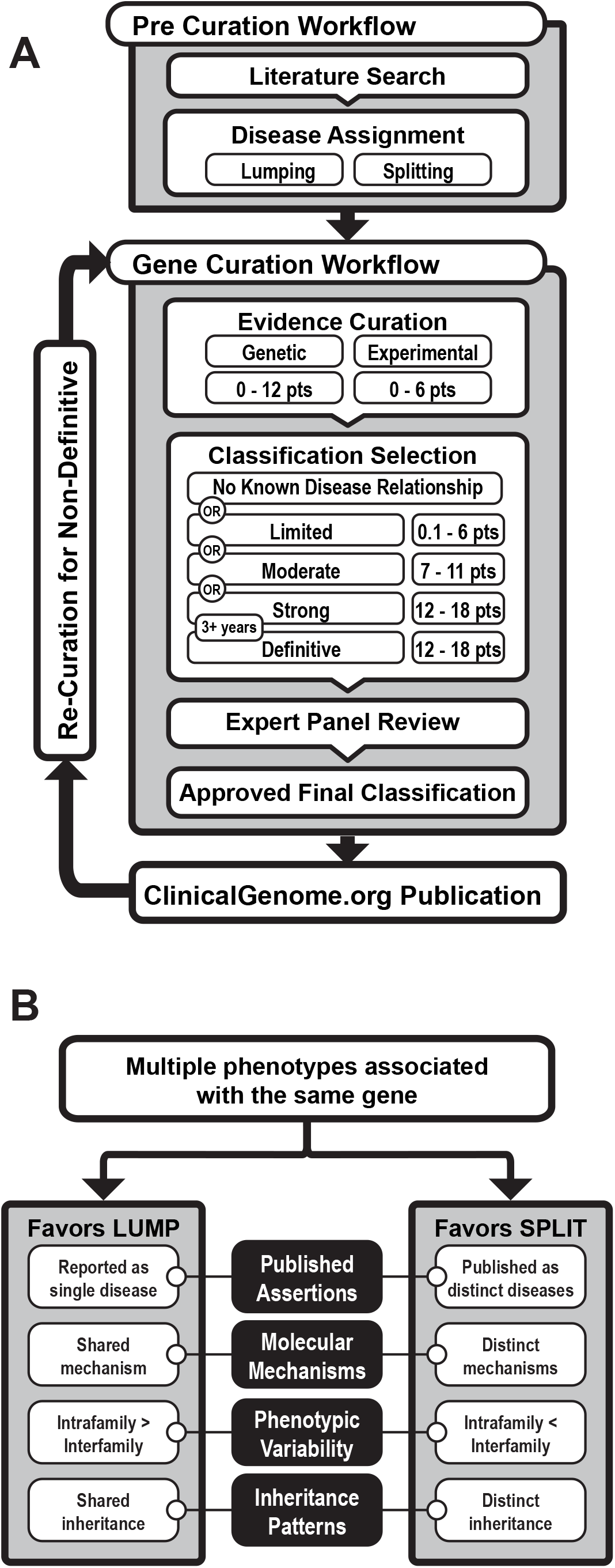
ClinGen gene-disease curation framework. **A)** Schematic overview of the ClinGen curation process. Adapted from ^14^. **B)** Decision framework for lumping or splitting of gene-disease relationships. Adapted from Thaxton *et al.* ^16^.

### Pre-curation, lumping, and splitting

Potential gene-disease relationships suitable for curation by the PIRD-GCEP were identified from Table 4 – Diseases of Immune Dysregulation, in the 2022 and 2024 IUIS IEI classifications ^3,15^ as well as GCEP members’ domain expertise. Several genes within the scope of the PIRD-GCEP are associated with multiple proposed gene-disease relationships. The decision to lump these varying entities into a larger aggregate gene-disease relationship or further split them into more granular gene-disease relationships was based on ClinGen’s previously described lumping and splitting guidelines ^16^.

### Gene curation

Primary gene curations were completed according to ClinGen SOP ^14^ versions 8-12, depending on the date of curation. Curations took place between July 2023 and April 2026 and were completed within the ClinGen Gene Curation Interface (GCI) ^17^. The curation process involves 3 major components: genetic evidence, experimental evidence, and final classification. The PIRD-GCEP also subsumed long-term management and recuration of gene-disease relationships from the AD-GCEP after completion of primary curations in 2025 ^13^.

Genetic evidence was evaluated from published case-level and case-control data using the ClinGen semiquantitative scoring framework (maximum 12 points). Starting scores for missense (+0.1) or predicted/proven null (+1.5) variants could be upgraded based on features such as accompanying functional/experimental data (+0.5) or *de novo* variant status (+0.5). Reasons for downgrading scores included the presence of a confounding alternative, disease-causing variant or uncertainty in the quality of patient data provided. Repeated instances of the same variant were downgraded or excluded if affected individuals shared a similar clinical and genetic background. Curators also recorded patient phenotypes (free text and HPO terms), demographics, genetic testing methods, and parental genetic testing data.

Experimental evidence (maximum 6 points) was evaluated across four evidence categories: functional evidence, functional alteration, model systems, and rescue studies. Functional evidence included studies supporting the normal biology of the gene product, such as tissue-specific expression, protein interactions, or disease-relevant biochemical activity. Functional alteration evidence included cellular or molecular studies demonstrating that disruption of gene or protein function produces disease-associated abnormalities. Model system evidence included animal or tissue-based models that recapitulate aspects of the human phenotype following gene disruption. Rescue evidence included studies in patients, patient-derived cells, or model systems showing reversal of the disease phenotype following restoration or correction of gene function. Each category included recommended scoring ranges and category-specific maximum point allocations.

Combined genetic and experimental scores were used to assign final gene-disease relationship classifications ranging from “limited” to “moderate” and “strong.” Relationships classified as “strong” were elevated to “definitive” if supported by replicated genetic evidence reported over a period of at least 3 years. The classification “disputed” was assigned in the presence of contradictory evidence, such as consistently low-scoring cases or previously reported variants later found to be too common in the general population or attributable to alternative genetic causes.

### Disease nomenclature

At the time of pre-curation, each GDR’s disease name was taken preferentially from MONDO, then OMIM, then GCEP expert consensus, using the first available source. This disease name was included as the formal disease name when the completed GDR curation was published to clinicalgenome.org. However, preferred disease names following a unified nomenclature determined by the PIRD-GCEP were suggested to MONDO after GDR curation (labeled in MONDO as “ClinGen Label”). If accepted, published ClinGen curations will be updated to reflect these new disease names.

### HPO compilation and similarity calculations

Proband HPO annotations were downloaded and compiled using ClinGen’s official Application Programming Interface (https://vci-gci-docs.clinicalgenome.org/vci-gci-docs/gci-help/gci-api). All ClinGen curation data up until April 2026 were included. Graph information content (GIC) ^18,19^ was used to quantify similarity between probands based on their associated HPO terms and was implemented using hpo3/1.3.1. Briefly, HPO information content (IC) weights rare phenotypic terms more heavily than common terms by calculating their inverse relative frequency within the larger HPO dataset. GIC is then calculated by obtaining the sum of IC for all shared (intersection) HPO terms and their common ancestor terms between the two probands and dividing it by the sum of IC for all HPO terms and their common ancestor terms in either of the two probands (union). The pairwise proband GIC similarity matrix for all probands was used as the input for UMAP reduction, implemented by umap/0.2.1.

### HPO phenotypic clustering

PIRD phenotypic clusters were determined based on co-occurrence of HPO terms. To control for curator-specific annotation bias, all HPO terms with a GIC similarity of > 0.9 were considered functionally equivalent. For a given GDR, the set of curated HPO terms was expanded to include all functionally equivalent HPO terms and this combined list was used to determine all possible HPO terms pairs for a given GDR. All resulting HPO pairs were tallied across GDRs and organized into an adjacency matrix used to construct an HPO co-occurrence graph, where nodes represent HPO terms and edges represent whether the terms co-occurred within a GDR (igraph/1.3.1 and ggraph/2.2.2, R). The resulting graph was then clustered using the Leiden algorithm to generate the PIRD phenotypic clusters (leiden/0.4.3.1, R).

### Software and data availability

All analyses were conducted in R/4.3.1 and Python/3.12.2. ClinGen data are publicly available at https://clinicalgenome.org/. All analytic code necessary to reproduce these results is available at https://github.com/BenSolomon/pirdGCEP.

## RESULTS

### Defining scope of the PIRD-GCEP

The PIRD-GCEP identified 51 genes from Table 4: Diseases of Immune Dysregulation in the 2022 IUIS IEI Classifications ^15^. Of these, 46 had not yet been scheduled for curation by other Immunology CDWG GCEPs. 18 genes were added based on updates to Table 4 in the 2024 IUIS IEI classifications ^3^, as well as 23 additional genes based on clinical domain expertise.

Several genes within the scope of the PIRD-GCEP were associated with multiple potentially distinct disease phenotypes within the published literature. As previously described ^16^, the decision to lump vs. split phenotypes associated with a single gene was based on whether phenotypic differences were sufficiently supported by distinct clinical, molecular, and inheritance patterns to justify separate disease entities rather than a phenotypic spectrum (**Figure 1B**). These criteria resulted in lumping of the immune and neuropathy phenotypes associated with *ITPR3,* while phenotypes associated with *STAT1* and *STAT5B* were split into multiple gene-disease relationships (GDRs) (**Table 1**).

**Table 1:**
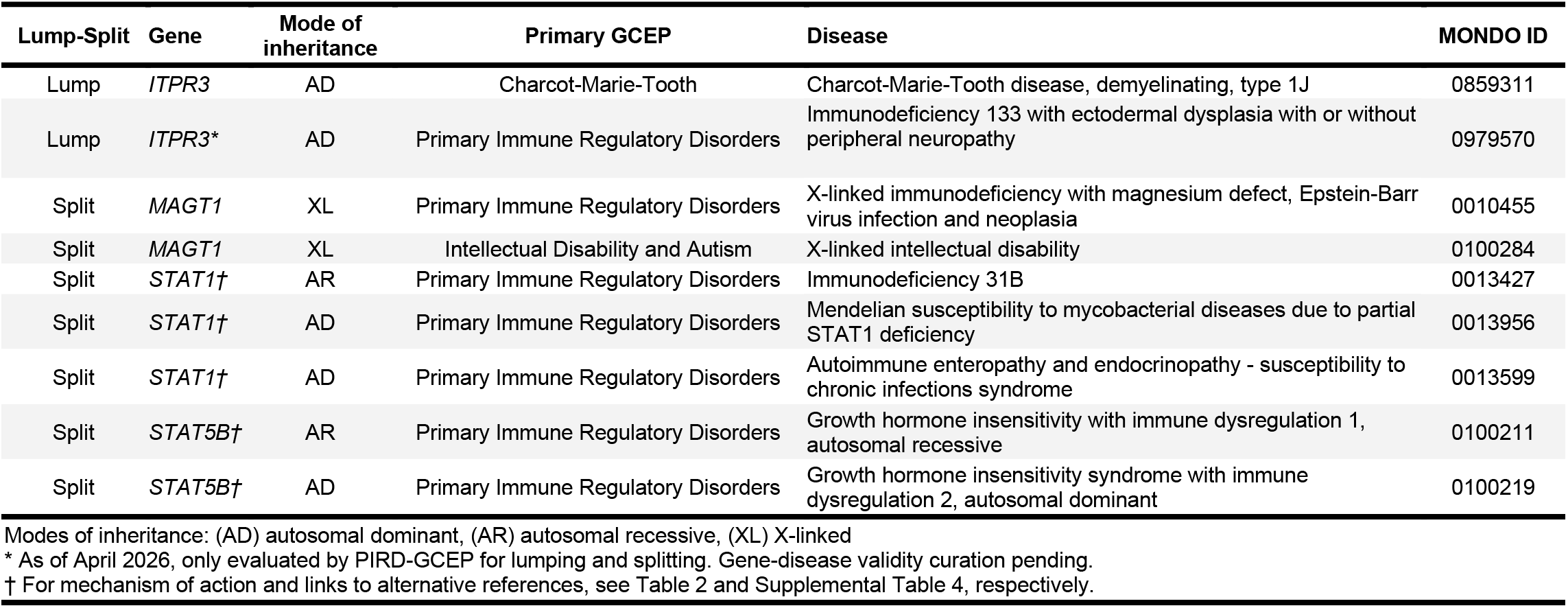
Lumped and split gene–disease relationships in genes causing PIRDs.

| Lump-Split | Gene | Mode of inheritance | Primary GCEP | Disease | MONDO ID |
| --- | --- | --- | --- | --- | --- |
| Lump | *ITPR3* | AD | Charcot-Marie-Tooth | Charcot-Marie-Tooth disease, demyelinating, type 1J | 0859311 |
| Lump | *ITPR3* * | AD | Primary Immune Regulatory Disorders | Immunodeficiency 133 with ectodermal dysplasia with or without peripheral neuropathy | 0979570 |
| Split | *MAGT1* | XL | Primary Immune Regulatory Disorders | X-linked immunodeficiency with magnesium defect, Epstein-Barr virus infection and neoplasia | 0010455 |
| Split | *MAGT1* | XL | Intellectual Disability and Autism | X-linked intellectual disability | 0100284 |
| Split | *STAT1*† | AR | Primary Immune Regulatory Disorders | Immunodeficiency 31B | 0013427 |
| Split | *STAT1*† | AD | Primary Immune Regulatory Disorders | Mendelian susceptibility to mycobacterial diseases due to partial STAT1 deficiency | 0013956 |
| Split | *STAT1*† | AD | Primary Immune Regulatory Disorders | Autoimmune enteropathy and endocrinopathy - susceptibility to chronic infections syndrome | 0013599 |
| Split | *STAT5B*† | AR | Primary Immune Regulatory Disorders | Growth hormone insensitivity with immune dysregulation 1, autosomal recessive | 0100211 |
| Split | *STAT5B*† | AD | Primary Immune Regulatory Disorders | Growth hormone insensitivity syndrome with immune dysregulation 2, autosomal dominant | 0100219 |

For example, *STAT1* was split into three separate GDRs. Two of these are characterized by susceptibility to mycobacterial infection but differ in both mode of inheritance and molecular mechanism. “Mendelian susceptibility to mycobacterial diseases due to partial STAT1 deficiency” (MONDO:0013956) is autosomal dominant due to confirmed dominant negative (DN) action of the variant *STAT1* protein, while “immunodeficiency 31B” (MONDO:0013427) is autosomal recessive and associated with complete or partial loss of function (LOF). Although the lumping and splitting guidelines support their separation into distinct GDRs based on differing inheritance patterns and molecular mechanisms, these two entities could nonetheless be considered mechanistically related, as both ultimately converge on impaired *STAT1* function as their shared end effect. The final GDR, “Autoimmune enteropathy and endocrinopathy - susceptibility to chronic infections syndrome” (MONDO:0013599), demonstrates autosomal dominant *STAT1* gain of function (GOF) and a distinct phenotype of susceptibility to fungal infection and autoimmunity.

### Gene curations

As of April 2026, the PIRD-GCEP has completed primary curations for 46 genes corresponding to 49 GDRs (**Table 2**). The median number of individuals curated per GDR was 7 (range: 1–16), reflecting both the inherent limits on the number of fully described cases available in the published literature and the capping of curations once a definitive classification was reached. For 9 additional GDRs with prominent non-immune features alongside an immune dysregulation phenotype, primary curation was conducted by other ClinGen GCEPs, with the PIRD-GCEP providing secondary curation to summarize evidence for the immunologic phenotype and assess concurrence with the primary GCEP’s final classification (**Supplemental Table 1**). However, the PIRD-GCEP did not contribute additional evidence scores or phenotypic annotations as part of these secondary curations. 11 additional genes from IUIS IEI Table 4 or with known immune regulatory phenotypes were curated by other Immunology CDWG GCEPs and did not require secondary curation by the PIRD-GCEP (**Supplemental Table 2**). Only those GDRs with a primary curation by the PIRD-GCEP are included in subsequent analyses. Currently, curations for 42 additional genes are planned (**Supplemental Table 3**).

**Table 2:**
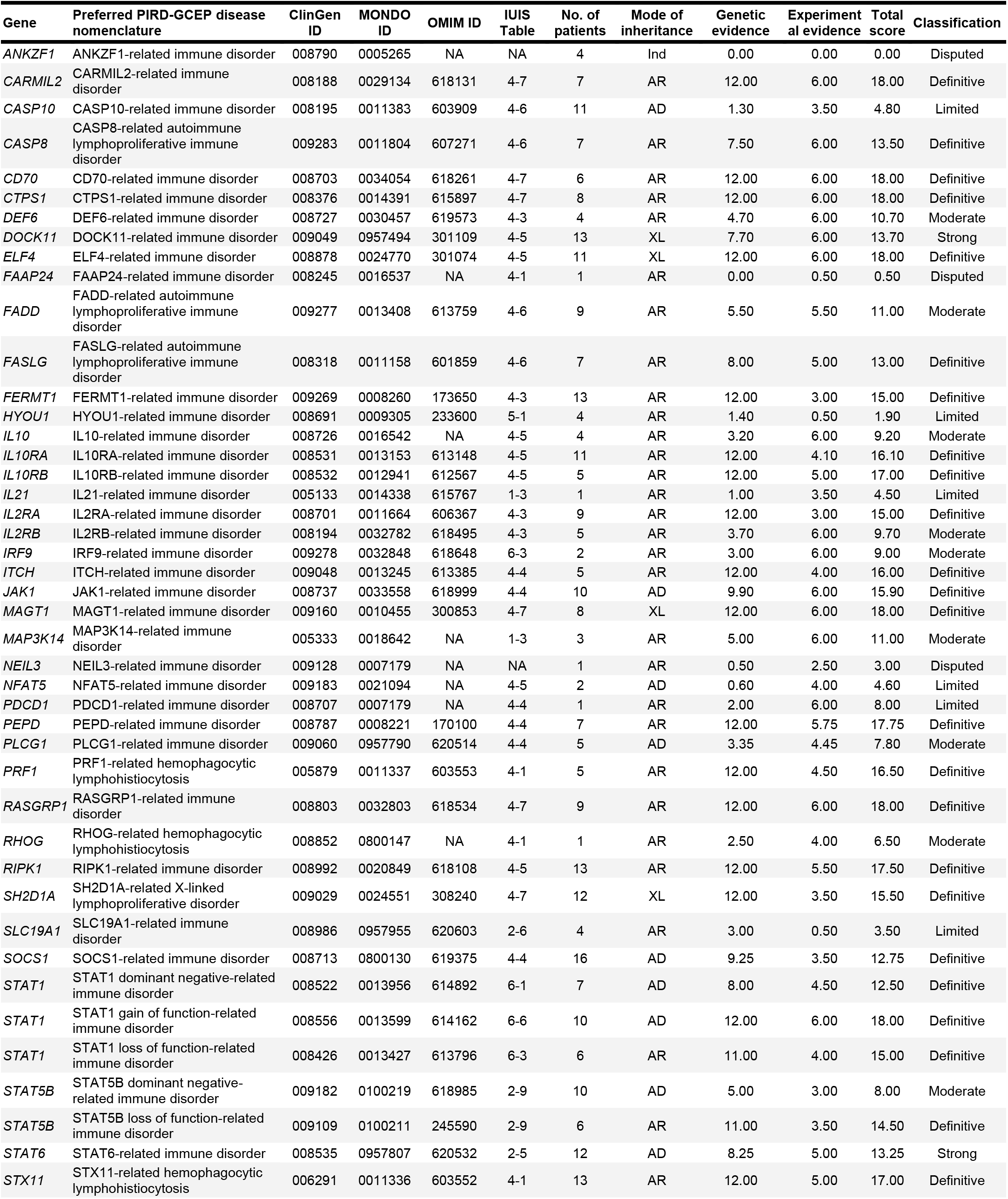
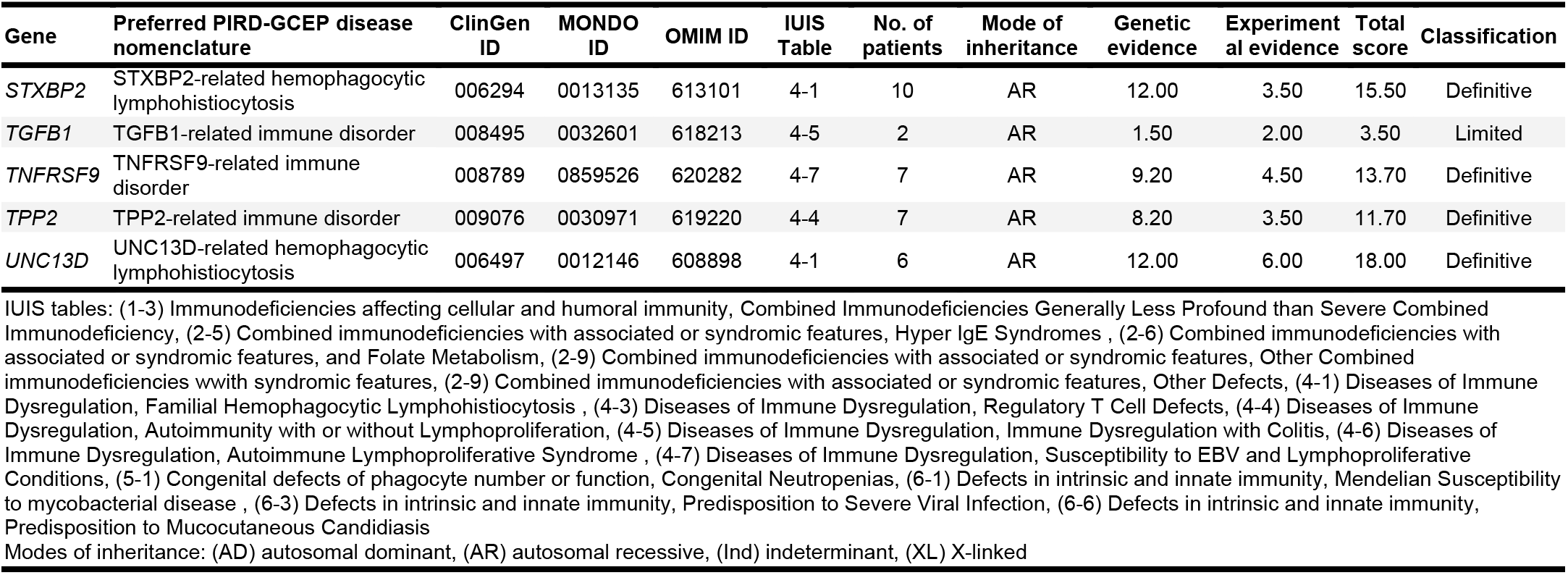
Classification of gene–disease relationships curated by the PIRD-GCEP using the ClinGen framework.

Evidence for each GDR was curated according to the SOP (**Figure 1A**). In brief, curated data is categorized as either genetic evidence, which correlates phenotype and genotype data from affected individuals, or experimental evidence, which includes laboratory-based data that supports a biological role for a gene in the pathogenesis of the observed condition. Genetic evidence can be assigned a maximum of 12 points, while experimental evidence can reach a maximum of 6 points. The combined genetic and experimental evidence score is the primary determinant of a GDR’s evidence classification, which can be reported as definitive, strong, moderate, limited, no known disease relationship, disputed, or refuted (**Figure 2A**).

**Figure 2:**
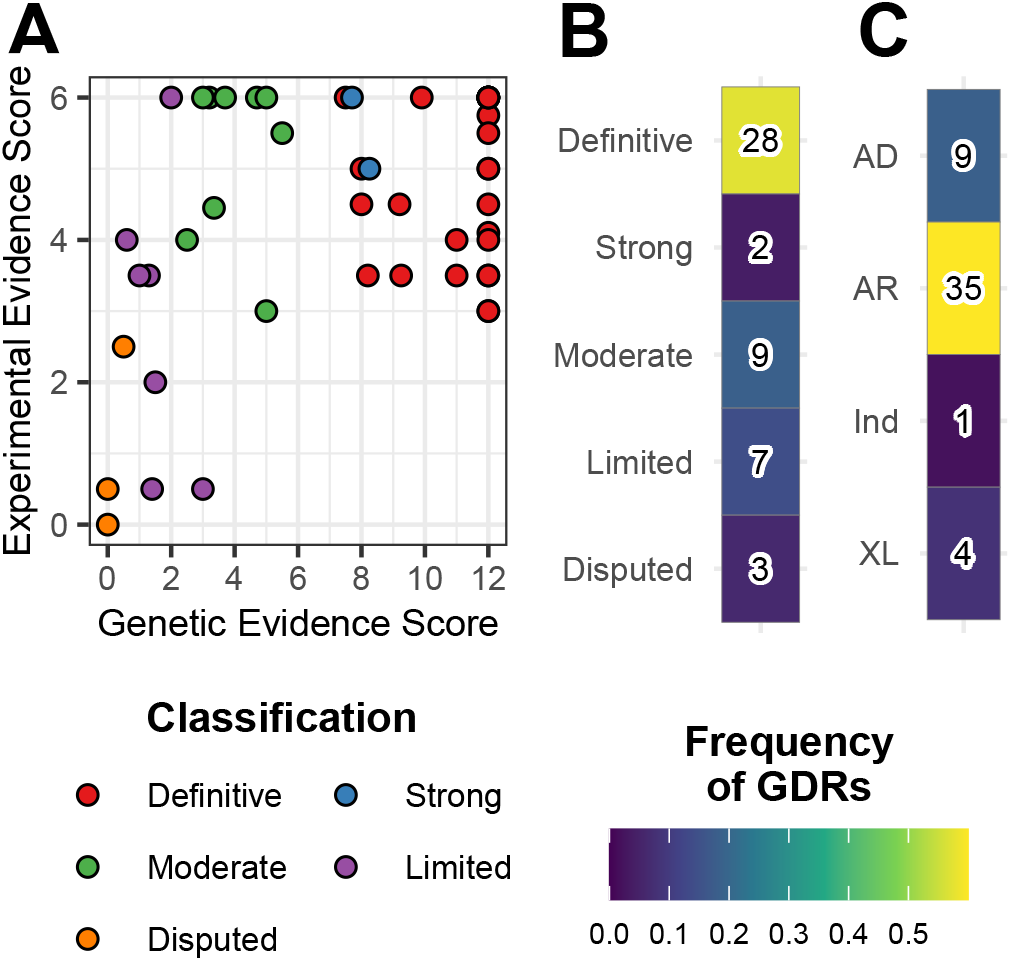
Evidence distribution and characteristics of PIRD gene-disease relationships. **A)** Contribution of genetic and experimental evidence to PIRD gene-disease curations. Points represent individual gene-disease curations, colored by their final evidence classification. **B)** Distribution of evidence classifications across all PIRD gene-disease curations. **C)** Distribution of modes of inheritance across all PIRD gene-disease curations. **B-C)** Colored by relative frequency across all gene-disease relationships and annotated with total number of curations. GDR = gene-disease relationship. AR = autosomal recessive. AD = autosomal dominant. XL = X-linked. Ind = indeterminate.

Of the completed PIRD-GCEP curations, 28 GDRs were categorized as definitive, 2 as strong, 9 as moderate, 7 as limited, and 3 as disputed (**Figure 2B**). The relative distribution of classifications is consistent with those obtained by other immunology GCEPs including the AD-GCEP ^13^ and the ClinGen Severe Combined Immune Deficiency-Combined Immune Deficiency (SCID-CID)-GCEP (**Supplemental Figure 1A**, Fisher exact p-value = 0.19).

35 conditions exhibited primarily autosomal recessive inheritance, 9 autosomal dominant, 4 X-linked, and 1 indeterminate (**Figure 2C**). The single indeterminate GDR was for a proposed *ANKZF1* early onset inflammatory bowel disease, based on a single case series of 4 patients; while the index patient had a biallelic variant with functional disruption of *ANKZF1* that could support an autosomal recessive mode of inheritance, the other three had alternative heterozygous or compound heterozygous variants without functional validation, precluding a definitive inheritance assignment ^20^. The overall inheritance patterns did differ significantly across all immunology GCEPs (**Supplemental Figure 1B**, Fisher exact p-value = 0.003), though this was primarily due to variation between genes curated by the AD-GCEP and SCID-CID-GCEP (adjusted Fisher exact p-value = 0.0001). In contrast, the inheritance patterns of PIRD-GCEP genes did not differ significantly from the other immunology GCEPs when adjusted for multiple comparisons.

Overall, the contribution of experimental and genetic evidence towards the final classification of a GDR was relatively balanced for all immunology GCEPs, though the AD-GCEP did demonstrate a small, but significant skewing in favor of experimental evidence (**Supplemental Figure 1C**). As expected, the curation of a larger number of affected individuals correlated with higher genetic evidence scores (**Supplemental Figure 1D**). Experimental evidence scores also increased with additional curated patients, but this was less significant than the effect on genetic evidence (linear model interaction p-value < 6×10^-13^).

### Representative curations

#### UNC13D – Definitive: Confirmation of a well-established disease relationship

Pathogenic variants in *UNC13D* are consistently among the most frequent genetic causes of fHLH ^21^. The role of *UNC13D* in fHLH pathogenesis has been known since 2003 ^22^ when it and its encoded protein, Munc13-4, were discovered based on identification in fHLH patients with intact perforin. As such, published patient data were readily available for curation and the GDR subsequently obtained the maximal number of genetic evidence points (12/12). As expected, this included many patients with predicted null variants leading to hemophagocytosis, hepatosplenomegaly, cytopenias, end-organ damage, and abnormal inflammatory markers and cytokines.

In addition, the initial discovery of *UNC13D* fit into an extensive body of evidence regarding the role of Munc family proteins in SNARE-mediated vesicle trafficking ^23^, leading to identification of Munc13-4’s role in CD8+ T cell and NK cell exocytosis of perforin- and granzyme-containing secretory lysosomes. Significant experimental data has since accumulated and was available for curation. This included evidence of a molecular association for Munc13-4 with lysosomes ^24^, the causative role of Munc13-4 deficiency in a mouse model of HLH ^25,26^, and the ability of exogenous *UNC13D* expression to rescue cytotoxicity in T cells from *UNC13D* deficient fHLH patients ^27^. As such, this gene-disease relationship achieved maximal experimental evidence points (6/6). Moreover, because multiple scorable cases were separated in time by more than 3 years, this gene-disease relationship was considered replicated over time and was able to receive the highest classification of “definitive.”

#### CASP10 – Limited: Standardized curation reveals insufficient evidence despite biological plausibility

Although less well characterized than cases caused by deleterious variants in *FAS* and *FASLG*, ALPS has also been attributed to variants in *CASP10*, with cases reported as early as 1999 ^28^. While 9 affected individuals with unique variants were identified for curation, relatively few points were assigned to these cases for reasons including relatively high allele frequencies (> 0.05) of reported variants in the general population (suggesting benign variation), the presence of confounding variants in other apoptosis pathway genes, and inconsistent phenotypic features. For example, one patient with a predicted null variant in *CASP10* presented with features more typical of Hyper IgE Syndrome, with the exception of hepatosplenomegaly ^29^.

Similarly, despite ample support for the role of *CASP10* in FAS-mediated apoptosis, only 3.5 points were assigned for experimental evidence. Caspase 10, encoded by *CASP10,* is recruited to both FAS and the TNF receptor by FADD upon ligand engagement ^30^. Variants that disrupt the active site of Caspase 10 interfere with both FASL- and TRAIL-mediated apoptosis ^31^, suggesting a DN mechanism. However, the same DN functional outcome was observed with both a reported rare variant as well as a common polymorphism ^32^. In contrast, recent data demonstrated that FAS-mediated apoptosis is unaffected by biallelic variants that completely abrogate *CASP10* expression ^33^. In addition, an ortholog of *CASP10* has been conspicuously lost in mice and rats ^34^, excluding the possibility of a common source of model organism-based experimental evidence, as well as supporting possible redundancy in caspases (e.g. Caspase 8) associated with FAS-mediated apoptosis. As a result, the final point total of 4.80/18 was assigned the classification of “limited.”

#### PDCD1 – Limited: Early but compelling evidence for a recently described relationship

The advent of immune checkpoint inhibitors (ICIs) has led to major advances in cancer therapy and, consequently, a substantial body of evidence describing their mechanisms of action and associated toxicities. Notably, these therapies can induce multisystem autoimmunity ^35^ and, counterintuitively, increase susceptibility to infection ^36,37^. Interestingly, PD-1-related ICIs are more likely to be associated with mycobacterial infection and reactivation compared to other ICIs ^38–40^. Consistent with these observations, a patient with a homozygous null variant in *PDCD1* (encoding PD-1) presented with type 1 diabetes, juvenile idiopathic arthritis, pulmonary autoimmunity, and peritoneal tuberculosis, ultimately succumbing to fatal autoimmunity ^41^.

Given the extensive literature on PD-1 inhibition, curation of this GDR by the PIRD-GCEP readily achieved the maximum score for experimental evidence (6/6). This included not only human data from checkpoint inhibitor studies but also supporting evidence from PD-1 deficient mouse models, which demonstrated both increased tuberculosis susceptibility ^42^ and spontaneous autoimmunity ^43^. However, because this relationship is supported by only a single reported patient, the genetic evidence score remained limited (2/12). Consequently, despite compelling mechanistic and experimental support, the overall classification of this GDR remains “limited.” However, given the consistency of existing evidence, the identification of additional cases would be expected to significantly strengthen this classification in future recurations.

### Harmonizing disease names among PIRDs

Currently, there is no unified nomenclature for naming PIRD diseases. As such, references to the same GDR often vary between different clinical resources, including the MONDO disease ontology, the Online Mendelian Inheritance in Man (OMIM) database, and the IUIS IEI classification tables. For instance, the PIRD associated with LOF variants in *ITCH* is referred to as “Syndromic multisystem autoimmune disease due to ITCH deficiency” in MONDO, “Autoimmune disease, multisystem, with facial dysmorphism” in OMIM, and “ITCH deficiency” in the IUIS IEI tables. Moreover, in the absence of clinical consensus for a gene-disease name within the literature, naming often follows the conventions of the databases in which they are found, leading to less recognizable and informative labels such as “immunodeficiency 64” for *RASGRP1* LOF variants or “inflammatory bowel disease 25” for *IL10RB* LOF variants.

Therefore, we sought to harmonize gene-disease labels among PIRDs (**Table 2**, **Supplemental Table 4**). Following the recommendations of ClinGen’s Disease Naming Advisory Committee ^44^, we adopted a dyadic naming strategy of the format “[gene]-related immune disorder.” In cases where a single gene was associated with multiple, split disease relationships, we further incorporated the presumed mechanism of the gene defect, such as “STAT1 LOF-related immune disorder.” Finally, for several long-established disease names, we retained the conventional name in the dyadic naming scheme, such as “autoimmune lymphoproliferative immune disorder,” “familial hemophagocytic lymphohistiocytosis”, and “X-linked lymphoproliferative disease.”

### Standardized phenotypic annotations

To standardize the representation of phenotypes in GDRs, ClinGen curators annotate patient histories and clinical data utilizing the Human Phenotype Ontology (HPO). This biological ontology is a vocabulary of over 17,000 terms arranged into a directed acyclic graph (DAG) ^45^. This representation allows for standardization of phenotypic descriptions and also enables subsequent evaluation of these HPO-annotated phenotypes to account for the similarity of related but distinct terms based on their proximity within the DAG structure. To date, curations completed by the PIRD-GCEP include 2131 total HPO annotations, averaging 8 HPO terms per patient case. 648 of the PIRD-GCEP HPO terms were unique, averaging 13 unique terms per GDR.

We utilized GIC similarity to quantify phenotypic concordance between individual patients based on their associated HPO annotations and the relationship between those HPO terms within the ontology. By accounting for HPO term similarity, GIC similarity reduces the impact of bias originating from annotator-specific HPO annotation practices. We then utilized pairwise GIC similarity between all patients as the basis for dimensional reduction to visualize the overall phenotypic structure of PIRD-GCEP GDRs.

We observed that the clustering of individual PIRD patients reflected broad phenotypic features. First, patients generally clustered according to the IUIS IEI tables and sub-tables of their condition. For example, patients corresponding to PIRD GDRs outside of IUIS IEI Table 4 formed independent clusters (**Figure 3A**). Similarly, patients corresponding to several IUIS IEI Table 4 sub-tables, such as those associated with fHLH, ALPS-like conditions, or regulatory T cell defects, formed discrete clusters (**Figure 3B**). Unsurprisingly, mapping specific phenotypic terms onto these representations showed concordance with expected patient clusters, such as between “hemophagocytosis” and the fHLH sub-table genes (**Figure 3C**) as well as “colitis” and the immune dysregulation with colitis sub-table genes (**Figure 3D**).

**Figure 3:**
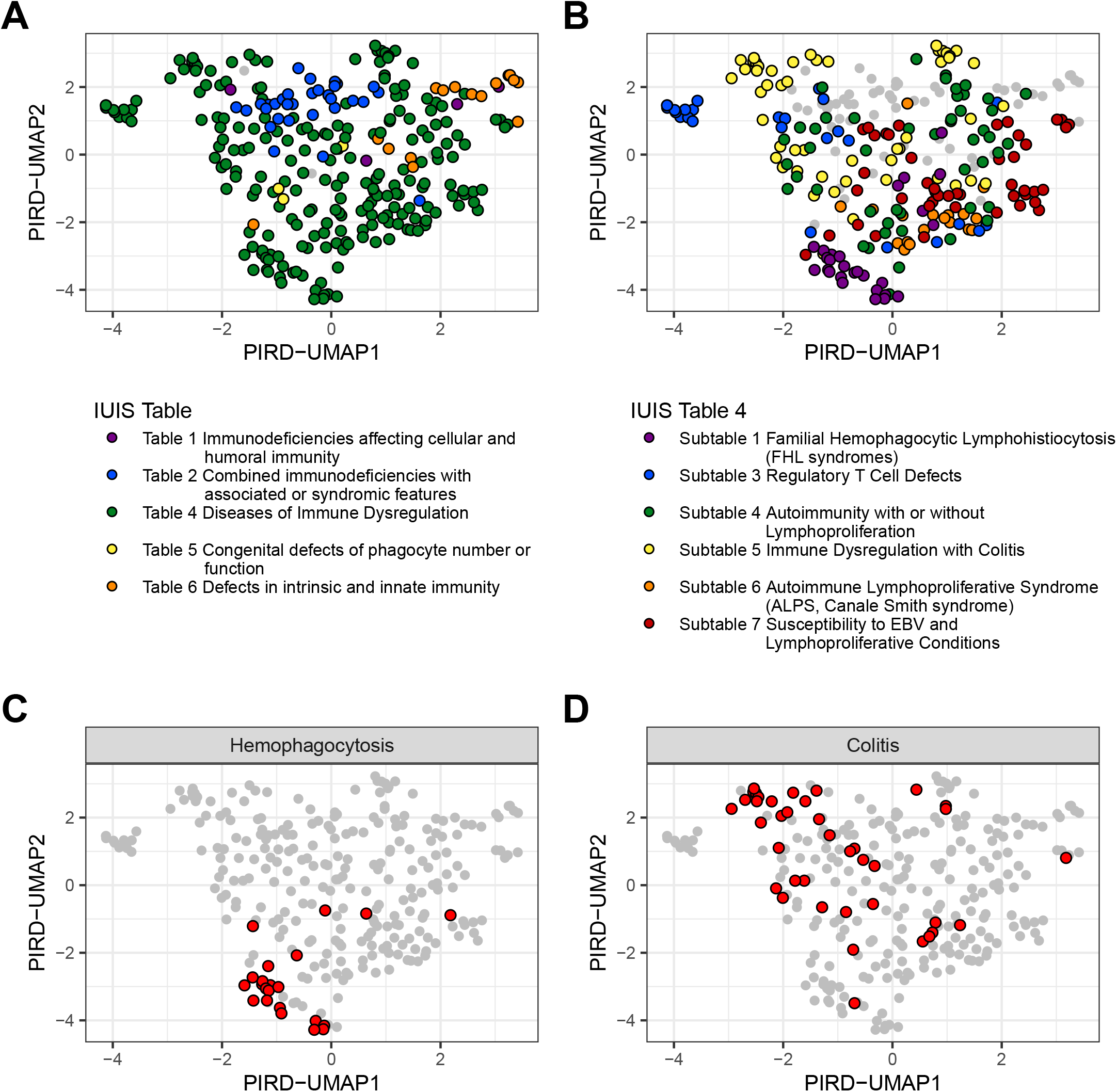
HPO-based phenotypic clustering of PIRD patients aligns with clinical classifications. Uniform manifold approximation and projection (UMAP) plots of proband similarity based on HPO annotations. Similarity values between all probands based on graph information content (GIC) similarity of HPO terms used as input for UMAP reduction of **A, C)** all immunology GCEP probands and **B, D)** PIRD-GCEP specific probands. Probands colored by the **A)** IUIS IEI table or **B)** IUIS IEI Table 4 subtable of the corresponding GDR. **C-D**) Probands highlighted if they were annotated with the corresponding HPO term.

### PIRDs can be characterized by 3 core phenotypic patterns

To objectively define phenotypic patterns across PIRDs, we constructed a network of co-occurring HPO terms and applied unsupervised clustering to identify distinct phenotypic groupings. We connected (i.e. edges) terms (i.e. nodes) that co-occurred within the same GDR and weighted both nodes and edges by their frequency across curated cases (**Figure 4A**). Because HPO annotations can vary depending on case description detail and ClinGen annotator preference, we accounted for potential bias arising from the use of closely related terms. For example, lymphadenopathy in ALPS may be annotated using a general term (“lymphadenopathy”), a more specific term (“generalized lymphadenopathy”), or multiple site-specific terms, depending on the level of detail available or annotator choices. Treating these as independent terms can fragment co-occurrence patterns and obscure true phenotypic relationships. To mitigate this, we used GIC similarity to identify highly related HPO terms and grouped them accordingly. Co-occurrence relationships were then extended across these similar terms, allowing phenotypic connections to be captured even when different but closely related terms were used. This approach reduces the impact of annotation variability and improves the robustness of inferred phenotypic clusters.

**Figure 4:**
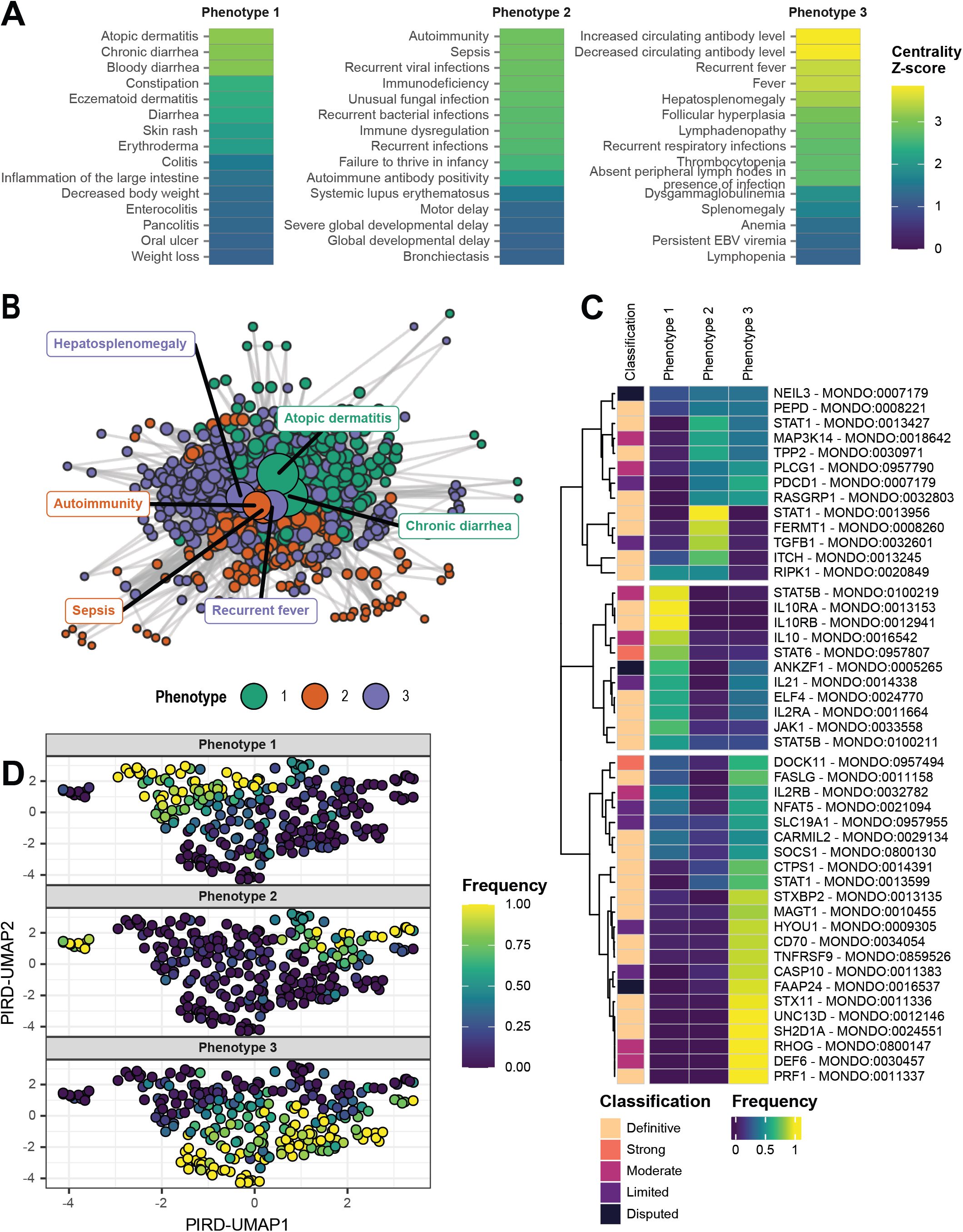
HPO co-occurrence reveals three core phenotypic modules in PIRDs. **A)** Top 15 HPO terms associated with each phenotype cluster as determined by centrality within the HPO co-occurrence graph. **B)** HPO co-occurrence graph. Nodes (points) represent individual HPO terms. Edges (lines) connect pairs of HPO terms co-occurring within the set of all terms annotated for a single GDR. Additional edges drawn to connect all pairs of highly similar HPO terms with a GIC similarity of > 0.9 relative to the curated terms. Size of nodes represents relative frequency of HPO terms among all PIRD-GCEP HPO annotations. Weight of edges represents relative frequency of HPO pair among all PIRD-GCEP HPO annotation pairs. Node colors represent phenotype group determined by Leiden clustering. Text boxes highlight representative HPO terms from each cluster. **C)** Relative frequency of HPO terms belonging to each of the three phenotypic clusters for each PIRD gene-disease relationship. **D)** UMAP projection from Figure 3 overlaid with relative frequency of HPO terms belonging to each of the 3 phenotypic clusters.

Unsupervised clustering of the resulting PIRD network identified three major phenotypic modules (**Figure 4B**). Phenotype 1 captures atopic and gastrointestinal inflammation, with enrichment for dermatitis, diarrhea, and colitis, reflecting barrier and immune regulatory dysfunction. Phenotype 3 is characterized by lymphoproliferation and systemic inflammation, including lymphadenopathy, hepatosplenomegaly, cytopenias, and fevers, consistent with dysregulated immune activation. In contrast, Phenotype 2 represents both a combination of immune deficiency and classic autoimmunity, including recurrent or unusual bacterial, viral, and fungal infections, bronchiectasis, lupus, and autoantibodies.

We then evaluated the relative enrichment of these phenotypic clusters across PIRD GDRs by assigning individual HPO annotations to their corresponding phenotypic clusters. Projecting these modules onto GDRs revealed strong and biologically consistent segregation (**Figure 4C**). All classic fHLH genes (*PRF1, UNC13D, STXBP2, STX11*) expressed Phenotype 3 almost exclusively. Given the role of EBV in fHLH pathogenesis, it was not surprising to observe similar phenotypic expression among other GDRs characterized by EBV susceptibility such as *SH2D1A*, *MAGT1,* and *TNFRSF9*. Conditions involving genes important for Treg (*IL10, IL10RA, IL10RB, STAT5B, IL2RA*) and Th2 (*STAT6*) cell function all showed disproportionate expression of Phenotype 1, likely due to prominence of enteropathy and atopy, respectively. Notably, these phenotypic clusters appropriately segregated the three split *STAT1*-associated monogenic conditions. Both LOF (MONDO:0013427) and DN (MONDO:0013956) *STAT1* conditions showed predominant expression of Phenotype 2 correlating with known mycobacterial susceptibility. In contrast, GOF *STAT1* (MONDO:0013599) was more strongly associated with Phenotype 1 due to presence of enteropathy.

Finally, projecting these phenotypic clusters onto individual patients revealed distinct patient-level clustering (**Figure 4D**). While these network-derived modules broadly aligned with IUIS IEI Table 4 sub-tables (**Figure 3B**), they demonstrated more discrete phenotypic clusters compared to expert-consensus classifications. For instance, the sharp segregation of Phenotype 3 among fHLH and EBV susceptibility disorders, and Phenotype 1 among Treg and Th2 defects, suggests these modules may represent more objectively defined subgroups based on convergent pathophysiology and similar approaches could define more granular PIRD endotypes capable of guiding disease management.

## DISCUSSION

Through systematic application of ClinGen’s standardized curation framework, we have established definitive or strong evidence for 30 of 49 curated PIRD GDRs, providing a robust foundation for clinical interpretation of genetic variants in these conditions. Notably, our curations incorporated data from over 340 patients and generated more than 2,000 standardized phenotypic annotations, enabling the first comprehensive, ontology-based characterization of the PIRD phenotypic landscape. Unsupervised clustering of co-occurring HPO terms revealed three distinct phenotypic modules that align with underlying pathophysiology, including lymphoproliferation and systemic inflammation, atopic and gastrointestinal inflammation, and combined immune deficiency with classic autoimmunity.

One promising application of this resource is the use of standardized, machine-readable phenotypic data to train predictive models capable of accelerating IEI recognition. IEIs remain substantially underdiagnosed, with reported delays of 3–7 years between symptom onset and diagnosis, contributing to increased morbidity and healthcare utilization ^46^. Automated detection models integrated within electronic health record (EHR) systems may help reduce this diagnostic delay. Several early approaches have shown promise but frequently rely on International Classification of Diseases (ICD) codes or claims data ^47–50^, which were designed primarily for medical billing purposes and often inadequately capture the complexity of rare disease phenotypes ^51,52^. In contrast, the use of HPO-based phenotypic representations for IEI detection remains comparatively limited but has yielded encouraging results. For example, Johnson et al. incorporated HPO terms, among several other parameters, into a common variable immunodeficiency (CVID) prediction model capable of identifying affected patients a median of 244 days prior to clinical diagnosis ^53^. Similarly, Alsaati et al. directly incorporated HPO terms into a machine learning model that identified some CVID patients from EHR data up to 10 years before clinical diagnosis ^54^.

Our phenotype-driven clustering of PIRDs may also reveal convergent pathophysiology not readily captured by traditional expert-derived classifications, with the potential to mechanistically inform clinical management. Knowledge graphs that integrate HPO terms with other biologic ontologies have proven powerful for such tasks. For example, Zhu et al. used this approach to identify novel drug-repurposing targets in rare disease ^55^, and Alsentzer et al. applied an HPO-inclusive knowledge graph to identify causative variants among Undiagnosed Diseases Network (UDN) patients, even in genes with no previously established gene-disease relationship ^56^. Building on these approaches, the PIRD phenotypic data and clustering described here could help extend established therapies from well-characterized PIRDs to rarer or newly recognized ones, as well as facilitate the identification and validation of novel PIRD GDRs.

One limitation of our study is that the goal of ClinGen curation is to establish authoritative support for GDRs rather than to comprehensively capture the full phenotypic spectrum associated with each disorder. Accordingly, once the maximum allowable genetic or experimental evidence score is achieved, additional evidence of that category is not required to be considered nor curated. As a result, phenotypic annotations for GDRs classified as strong or definitive may reflect only a subset of reported cases, potentially introducing sampling bias into comparative analyses of phenotypes across PIRDs. Similarly, ClinGen curations may be susceptible to ascertainment bias, as the phenotype of a GDR is likely to be defined initially by its most severe presentations, with milder phenotypes recognized only later as the spectrum expands ^57^, though the ClinGen lumping and splitting guidelines seek to compensate for such situations by allowing for a new GDR to be curated. In contrast, resources such as GenIA make a different trade off, prioritizing broader aggregation of published IEI patient data, including standardized HPO annotations, over classifying gene-disease validity ^58^. This yields extensive curated patient data but does not assess the quality of that data and is limited to fewer GDRs.

Although HPO terms provide a standardized and computational framework for representing clinical phenotypes, their use also introduces important limitations. Most notably, HPO-based analyses are susceptible to author- and annotator-dependent bias. Curators are limited to the details included in source publications and thus curated phenotypes are only as detailed as the phenotypes described by the original authors. Moreover, as discussed previously, overlapping or closely related HPO terms may lead different curators to reasonably assign different annotations to the same clinical description ^58^. If the hierarchical relationships between HPO terms are not considered, this variability can distort downstream phenotypic analyses. In addition, curators may differ in the level of granularity used during annotation, ranging from assignment of a single broad descriptor (e.g., generalized lymphadenopathy) to multiple site-specific terms describing the same clinical finding. Automated HPO annotation approaches using natural language processing and large language models may represent a strategy to reduce curator-specific variability, though validation and large-scale implementation of existing models remain limited ^59–61^.

Limitations in HPO completeness and structure also represent important considerations. Despite containing more than 17,000 terms and being designed with rare disease applications in mind, highly specialized findings, particularly from niche immunologic diagnostic assays, may lack corresponding HPO terms. In addition, clinically related phenotypes may not always be optimally connected within the ontology structure, limiting the ability of graph-based similarity methods to capture semantic relationships between terms. For example, Haimel *et al.* demonstrated that expansion and restructuring of immunology-specific HPO terms significantly improved computational matching of IEI phenotypes to their corresponding diagnoses ^62^. Importantly, HPO solicits feedback from the clinical and research community, selectively integrating suggestions into revisions of the ontology.

In summary, we have leveraged ClinGen’s systematic curation framework to establish a comprehensive landscape of PIRD GDRs and their associated phenotypes. By integrating genetic evidence, experimental data, and standardized HPO annotations across 49 GDRs and over 340 patients, we have strengthened the evidence base for clinical variant interpretation and revealed three biologically coherent phenotypic modules that transcend traditional classification schemes. The standardized, machine-readable nature of these curations allows them to serve as a valuable resource for diagnostic prediction models, therapeutic stratification, and mechanistic insight into immune homeostasis.

## Data Availability

ClinGen curations are updated periodically, to find the most current information please visit clinicalgenome.org. Information for the PIRD-GCEP can be found at https://clinicalgenome.org/affiliation/40145/. All analytic code necessary to reproduce these results is available at https://github.com/BenSolomon/pirdGCEP.

https://www.clinicalgenome.org/

https://github.com/BenSolomon/pirdGCEP

## ACKNOWLEDGEMENTS

We are grateful to the University of North Carolina ClinGen Core for their guidance and support of the Immunology CDWG, AD-GCEP and PIRD-GCEP, including Dr. Jonathan Berg, Dr. Courtney Thaxton, Marwa Elnagheeb, Carolyn McCormick, Christina Gutierrez Ford, Haley Garrett, and Shannon Kelly. The Immunology CDWG and the PIRD-GCEP specifically acknowledge Dr. Courtney Thaxton for the continuous and broad support provided as the liaison from ClinGen to the Immunology CDWG. We also acknowledge the other co-chair of the Immunology CDWG, Dr. Ivan Chinn, for their support. We also thank Pharming for their partnership with ClinGen curation activities through the Immunology CDWG.

## ABBREVIATIONS

AD: Autosomal dominant
AD-GCEP: Antibody Deficiency GCEP
ALPS: Autoimmune lymphoproliferative syndrome
AR: Autosomal recessive
CID: Combined immune deficiency
CDWG: Clinical Domain Working Group
ClinGen: Clinical Genome Resource
DN: Dominant negative
GCEP: Gene Curation Expert Panel
GDR: Gene-disease relationship
GIC: Graph information content
GOF: Gain of function
fHLH: Familial hemophagocytic lymphohistiocytosis
HPO: Human Phenotype Ontology
IEI: Inborn error of immunity
IUIS: International Union of Immunological Societies
LOF: Loss of function
MONDO: Mondo Disease Ontology
OMIM: Online Mendelian Inheritance in Man
PIRD: Primary immune regulatory disorder
SCID: Severe combined immune deficiency
XL: X-linked

## SUPPLEMENTAL FIGURE LEGENDS

**Supplemental Figure 1:**
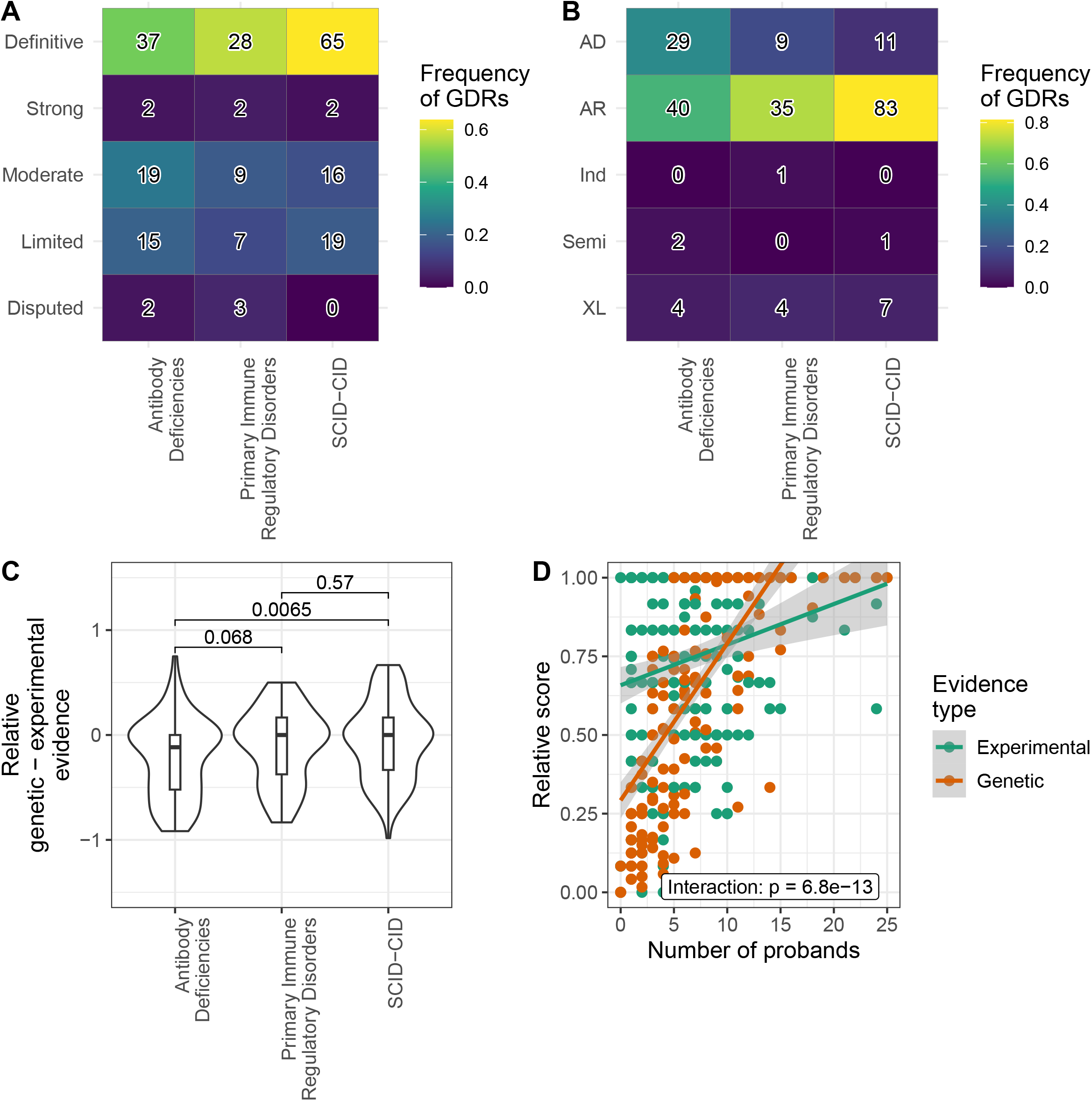
Comparative gene-disease validity across all ClinGen immunology GCEPs. **A)** Distribution of evidence classifications and **B)** modes of inheritance for GDRs across all immunology gene-disease curations, separated by GCEP. Colored by relative frequency of classification level across all GDRs and annotated with total number of curations. **C)** Relative weight of genetic vs. experimental evidence in determining final classification for GDRs across each immunology GCEP. Relative genetic and experimental evidence determined by standardizing component scores as a percentage of total possible points (12 for genetic evidence, 6 for experimental evidence). Difference between relative genetic and experimental evidence shown. **D)** Impact of number of curated probands from published literature on genetic and experimental point totals. Genetic and experimental evidence evaluated as relative point totals as described for **C)**. P-value reflects that associated with interaction term between number of probands and evidence type in linear model.

**Supplemental Table 1:** Gene-disease relationships with secondary curation performed by PIRD-GCEP.

| Gene | Disease | Primary GCEP | ClinGen ID | MONDO ID | OMIM ID | IUIS Table | Mode of inheritance | Classification |
| --- | --- | --- | --- | --- | --- | --- | --- | --- |
| *AP3B1* | Hermansky-Pudlak syndrome 2 | Hemostasis Thrombosis | 004146 | 0011997 | 608233 | 4-2 | AR | Definitive |
| *AP3D1* | Hermansky-Pudlak syndrome 10 | Hemostasis Thrombosis | 004147 | 0014885 | 617050 | 4-2 | AR | Moderate |
| *CD27* | Lymphoproliferative syndrome 2 | Childhood, Adolescent and Young Adult Cancer Predisposition | 009403 | 0014054 | 615122 | 4-7 | AR | Definitive |
| *FAS* | FAS-related autoimmune lymphoproliferative immune disorder | Childhood, Adolescent and Young Adult Cancer Predisposition | 009322 | 1060194 | 601859 | 4-6 | Semi | Definitive |
| *FOXP3* | Immune dysregulation-polyendocrinopathy-enteropathy-X-linked syndrome | Prenatal | 008198 | 0010580 | 304790 | 4-3 | XL | Definitive |
| *LYST* | Chediak-Higashi syndrome | Hemostasis Thrombosis | 005310 | 0008963 | 214500 | 4-2 | AR | Definitive |
| *PRKCD* | Systemic lupus erythematosus | Monogenic Systemic and Incomplete Lupus Erythematosus | 008725 | 0007915 | 615559 | 4-7 | AR | Definitive |
| *RAB27A* | Griscelli syndrome type 2 | Syndromic Disorders | 005943 | 0011872 | 607624 | 4-2 | AR | Definitive |
| *SLC7A7* | Lysinuric protein intolerance | Aminoacidopathy | 006201 | 0009109 | 222700 | 4-1 | AR | Definitive |

IUIS tables: (4-1) Diseases of Immune Dysregulation, Familial Hemophagocytic Lymphohistiocytosis , (4-2) Diseases of Immune Dysregulation, FHL Syndromes with Hypopigmentation, (4-3) Diseases of Immune Dysregulation, Regulatory T Cell Defects, (4-6) Diseases of Immune Dysregulation, Autoimmune Lymphoproliferative Syndrome , (4-7) Diseases of Immune Dysregulation, Susceptibility to EBV and Lymphoproliferative Conditions
Modes of inheritance: (AR) autosomal recessive, (Semi) semidominant, (XL) X-linked
TBD: To be determined

**Supplemental Table 2:**
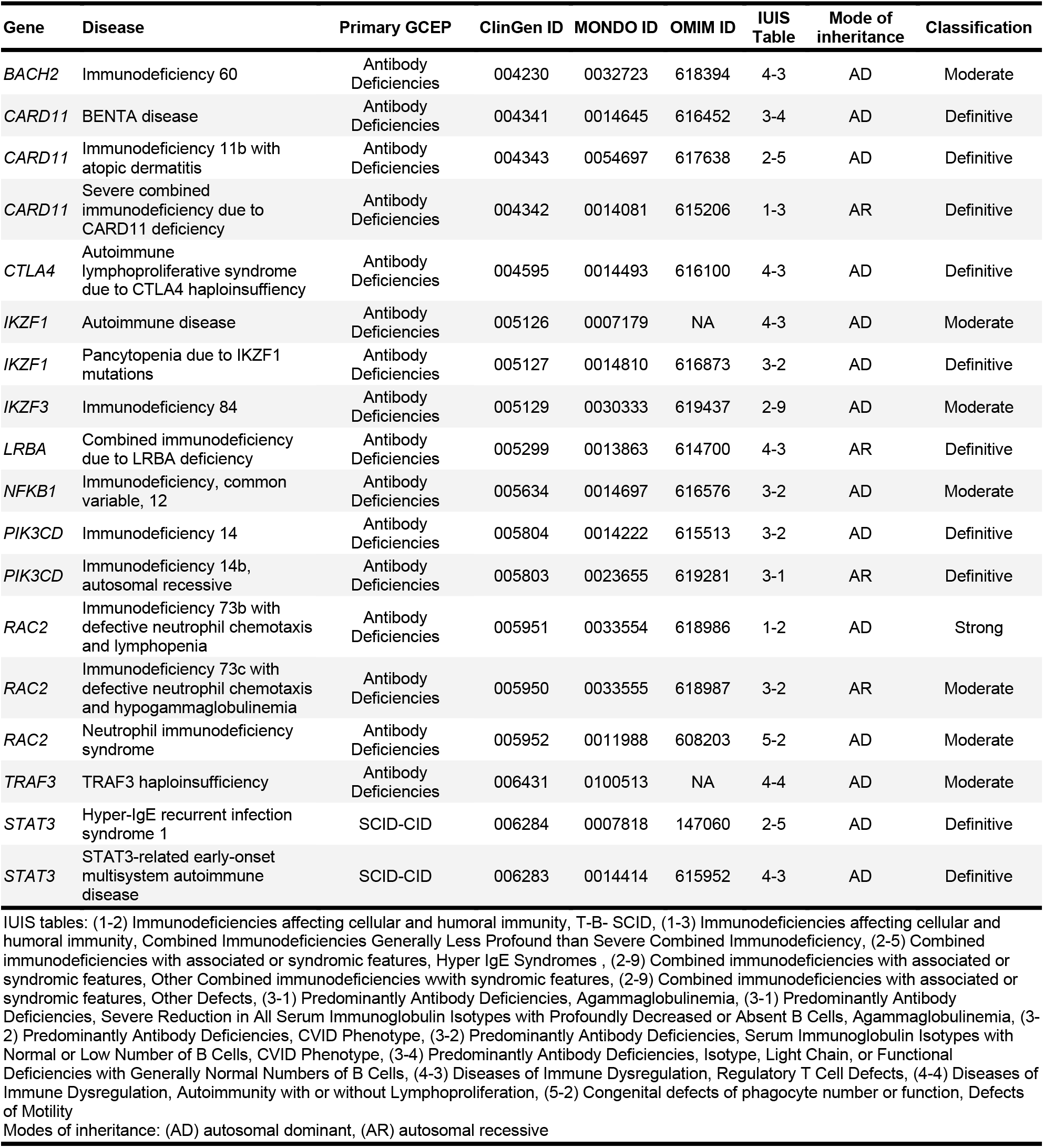
Immune regulatory gene–disease relationships curated by non-PIRD GCEPs.

| Gene | Disease | Primary GCEP | ClinGen ID | MONDO ID | OMIM ID | IUIS Table | Mode of inheritance | Classification |
| --- | --- | --- | --- | --- | --- | --- | --- | --- |
| *BACH2* | Immunodeficiency 60 | Antibody Deficiencies | 004230 | 0032723 | 618394 | 4-3 | AD | Moderate |
| *CARD11* | BENTA disease | Antibody Deficiencies | 004341 | 0014645 | 616452 | 3-4 | AD | Definitive |
| *CARD11* | Immunodeficiency 11b with atopic dermatitis | Antibody Deficiencies | 004343 | 0054697 | 617638 | 2-5 | AD | Definitive |
| *CARD11* | Severe combined immunodeficiency due to CARD11 deficiency | Antibody Deficiencies | 004342 | 0014081 | 615206 | 1-3 | AR | Definitive |
| *CTLA4* | Autoimmune lymphoproliferative syndrome due to CTLA4 haploinsufficiency | Antibody Deficiencies | 004595 | 0014493 | 616100 | 4-3 | AD | Definitive |
| *IKZF1* | Autoimmune disease | Antibody Deficiencies | 005126 | 0007179 | NA | 4-3 | AD | Moderate |
| *IKZF1* | Pancytopenia due to IKZF1 mutations | Antibody Deficiencies | 005127 | 0014810 | 616873 | 3-2 | AD | Definitive |
| *IKZF3* | Immunodeficiency 84 | Antibody Deficiencies | 005129 | 0030333 | 619437 | 2-9 | AD | Moderate |
| *LRBA* | Combined immunodeficiency due to LRBA deficiency | Antibody Deficiencies | 005299 | 0013863 | 614700 | 4-3 | AR | Definitive |
| *NFKB1* | Immunodeficiency, common variable, 12 | Antibody Deficiencies | 005634 | 0014697 | 616576 | 3-2 | AD | Moderate |
| *PIK3CD* | Immunodeficiency 14 | Antibody Deficiencies | 005804 | 0014222 | 615513 | 3-2 | AD | Definitive |
| *PIK3CD* | Immunodeficiency 14b, autosomal recessive | Antibody Deficiencies | 005803 | 0023655 | 619281 | 3-1 | AR | Definitive |
| *RAC2* | Immunodeficiency 73b with defective neutrophil chemotaxis and lymphopenia | Antibody Deficiencies | 005951 | 0033554 | 618986 | 1-2 | AD | Strong |
| *RAC2* | Immunodeficiency 73c with defective neutrophil chemotaxis and hypogammaglobulinemia | Antibody Deficiencies | 005950 | 0033555 | 618987 | 3-2 | AR | Moderate |
| *RAC2* | Neutrophil immunodeficiency syndrome | Antibody Deficiencies | 005952 | 0011988 | 608203 | 5-2 | AD | Moderate |
| *TRAF3* | TRAF3 haploinsufficiency | Antibody Deficiencies | 006431 | 0100513 | NA | 4-4 | AD | Moderate |
| *STAT3* | Hyper-IgE recurrent infection syndrome 1 | SCID-CID | 006284 | 0007818 | 147060 | 2-5 | AD | Definitive |
| *STAT3* | STAT3-related early-onset multisystem autoimmune disease | SCID-CID | 006283 | 0014414 | 615952 | 4-3 | AD | Definitive |

IUIS tables: (1-2) Immunodeficiencies affecting cellular and humoral immunity, T-B- SCID, (1-3) Immunodeficiencies affecting cellular and humoral immunity, Combined Immunodeficiencies Generally Less Profound than Severe Combined Immunodeficiency, (2-5) Combined immunodeficiencies with associated or syndromic features, Hyper IgE Syndromes, (2-9) Combined immunodeficiencies with associated or syndromic features, Other Combined immunodeficiencies with syndromic features, (2-9) Combined immunodeficiencies with associated or syndromic features, Other Defects, (3-1) Predominantly Antibody Deficiencies, Agammaglobulinemia, (3-1) Predominantly Antibody Deficiencies, Severe Reduction in All Serum Immunoglobulin Isotypes with Profoundly Decreased or Absent B Cells, Agammaglobulinemia, (3-2) Predominantly Antibody Deficiencies, CVID Phenotype, (3-2) Predominantly Antibody Deficiencies, Serum Immunoglobulin Isotypes with Normal or Low Number of B Cells, CVID Phenotype, (3-4) Predominantly Antibody Deficiencies, Isotype, Light Chain, or Functional Deficiencies with Generally Normal Numbers of B Cells, (4-3) Diseases of Immune Dysregulation, Regulatory T Cell Defects, (4-4) Diseases of Immune Dysregulation, Autoimmunity with or without Lymphoproliferation, (5-2) Congenital defects of phagocyte number or function, Defects of Motility
Modes of inheritance: (AD) autosomal dominant, (AR) autosomal recessive

**Supplemental Table 3:** Pending gene-disease curations by PIRD-GCEP.

| Gene | Disease | IUIS Table | MONDO ID | OMIM ID | PMID | Reference |
| --- | --- | --- | --- | --- | --- | --- |
| *ADAM17* | Inflammatory skin and bowel disease, neonatal, 1 | 7-3 | 0013693 | 614328 | 22010916 | 1 |
| *AGR2* | Respiratory infections, recurrent, and failure to thrive with or without diarrhea | NA | 0859370 | 620233 | 34952832 | 2 |
| *AIRE* | Autoimmune polyendocrine syndrome type 1 | 4-4 | 0009411 | 240300 | 35521792 | 3 |
| *AMFR* | To be determined | NA | NA | NA | 38277122 | 4 |
| *ARPC5* | Immunodeficiency 113 with autoimmunity and autoinflammation | 4-4 | 0957920 | 620565 | 37349293 | 5 |
| *CBLB* | Autoimmune disease, multisystem, infantile-onset, 3 | 4-4 | 0957388 | 620430 | 36006710 | 6 |
| *CD274* | Autoimmune disease, multisystem, infantile-onset, 5 | 4-4 | 0979235 | 621235 | 38634869 | 7 |
| *CD48* | To be determined | NA | NA | NA | 41984595 | 8 |
| *CDC42* | To be determined | 7-1 | NA | NA | 29394990 | 9 |
| *CEBPE* | Specific granule deficiency | 5-1 | 0009506 | 245480 | 758416 | 10 |
| *CEBPE* | CEBPE-associated autoinflammation-immunodeficiency-neutrophil dysfunction syndrome | 5-2 | 0035437 | 260570 | 31201888 | 11 |
| *DPP9* | Hatipoglu immunodeficiency syndrome | 4-1 | 0957229 | 620331 | 36112693 | 12 |
| *EIF2AK3* | Wolcott-Rallison syndrome | NA | 0009192 | 226980 | 40441422 | 13 |
| *FGL2* | To be determined | NA | NA | NA | 36243222 | 14 |
| *GIMAP6* | To be determined | 4-2 | NA | NA | 33328581 | 15 |
| *GNAI2* | To be determined | NA | NA | NA | 39298586 | 16 |
| *IKZF1* | To be determined | 1-3 | NA | NA | 27939403 | 17 |
| *IL27RA* | Immunodeficiency 134 (Epstein-Barr virus-specific) | 4-7 | 0980729 | 621405 | 38509369 | 18 |
| *ITGAV* | Immune dysregulation, neurodevelopmental defects, and colitis | NA | 0980702 | 621375 | 39526957 | 19 |
| *ITPR3* | Immunodeficiency 133 with ectodermal dysplasia with or without peripheral neuropathy | 2-8 | 0979570 | 621254 | 36302985 | 20 |
| *LACC1* | Juvenile arthritis due to defect in LACC1 | 4-4 | 0032920 | 618795 | 25220867 | 21 |
| *LY96* | To be determined | 6-7 | NA | NA | 36462957 | 22 |
| *NBEAL2* | Gray platelet syndrome | 4-3 | 0007686 | 139090 | 20709904 | 23 |
| *NCKAP1L* | Immunodeficiency 72 with autoinflammation | NA | 0033551 | 618982 | 32647003 | 24 |
| *NFATC1* | NFATC1-related combined immunodeficiency | 1-3 | 1010174 | NA | 37249233 | 25 |
| *NFATC2* | Joint contractures, osteochondromas, and B-cell lymphoma | 4-4 | 0859369 | 620232 | 35789258 | 26 |
| *RC3H1* | Hemophagocytic lymphohistiocytosis, familial, 6 | NA | 0033557 | 618998 | 31636267 | 27 |
| *RHBDF2* | To be determined | 4-5 | NA | NA | 34937930 | 28 |
| *RNU4ATAC* | To be determined | 2-4 | NA | NA | 41864208 | 29 |
| *RNU6ATAC* | To be determined | NA | NA | NA | 41808409 | 30 |
| *SH2B3* | SH2B3-related immune system disorder | 4-4 | 1060195 | NA | 40481232 | 31 |
| *SH3GL1* | To be determined | NA | NA | NA | 39499372 | 32 |
| *SRP19* | To be determined | 5-1 | NA | NA | 36223592 | 33 |
| *SRPRA* | To be determined | 5-1 | NA | NA | 36223592 | 34 |
| *STAT4* | Disabling pansclerotic morphea of childhood | 7-1 | 0957497 | 620443 | 37256972 | 35 |
| *TLR1* | To be determined | NA | NA | NA | 37479888 | 36 |
| *TLR1* | Inflammatory bowel disease 32 | NA | NA | 621601 | 42048460 | 37 |
| *TLR4* | To be determined | 6-7 | NA | NA | 36462956 | 38 |
| *TLR7* | Systemic lupus erythematosus 17 | 4-4 | 0859083 | 301080 | 35477763 | 39 |
| *TLR7* | Immunodeficiency 74, COVID-19-related, X-linked | 6-3 | 0026767 | 301051 | 35708626 | 40 |
| *TNFAIP3* | A20 haploinsufficiency | 7-3 | 0100222 | 616744 | 26642243 | 41 |
| *TNFSF9* | Immunodeficiency 109 with lymphoproliferation | 4-7 | 0859526 | 620282 | 30872117 | 42 |
| *TOM1* | Immunodeficiency 85 and autoimmunity | NA | NA | 619510 | 31263572 | 43 |
| *UNC93B1* | Herpes simplex encephalitis, susceptibility to, 1 | 6-4 | 0024563 | 610551 | 16973841 | 44 |
| *XIAP* | X-linked lymphoproliferative disease due to XIAP deficiency | 4-7 | 0010385 | 300635 | 17080092 | 45 |

**Supplemental Table 4:**
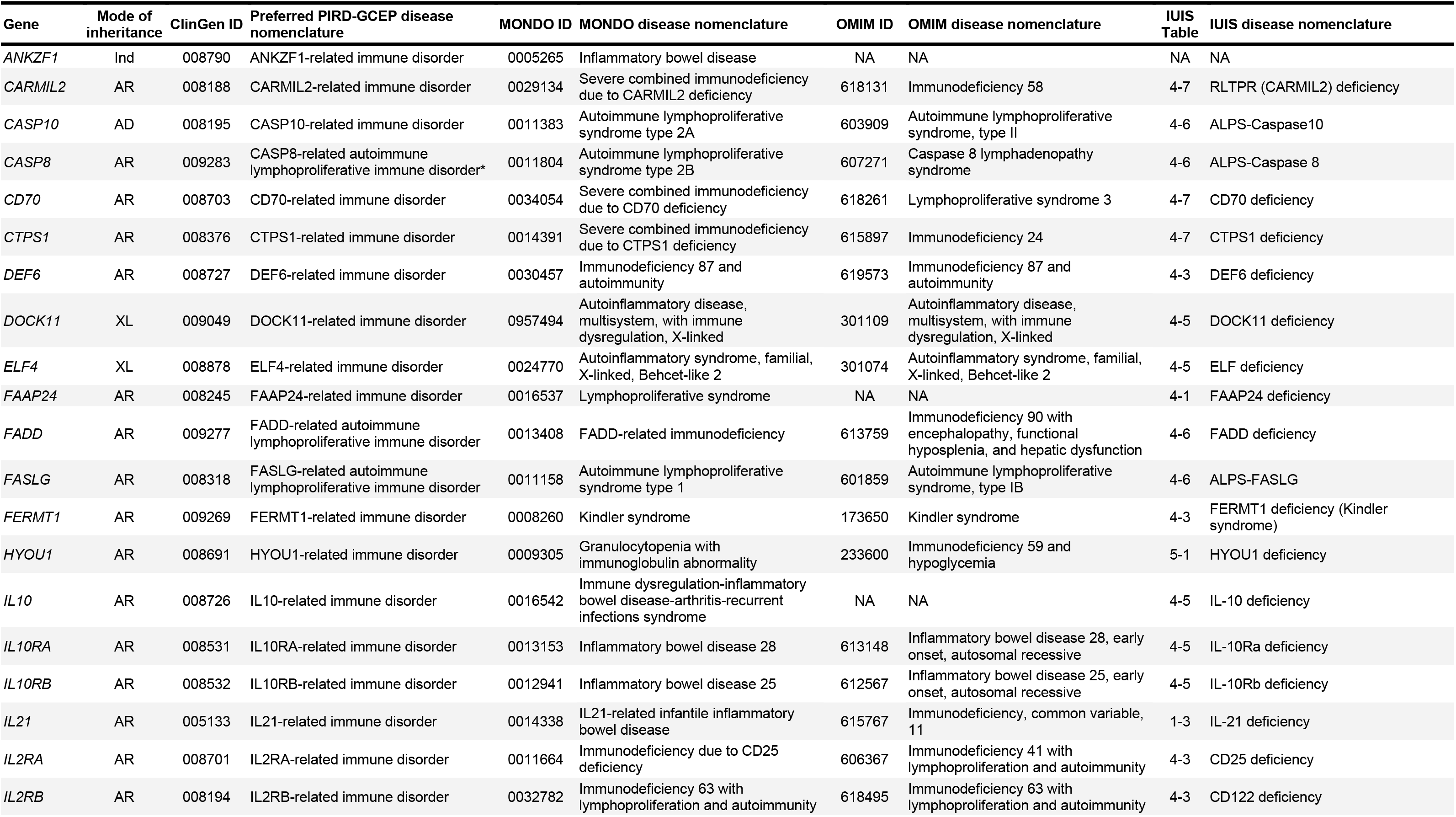
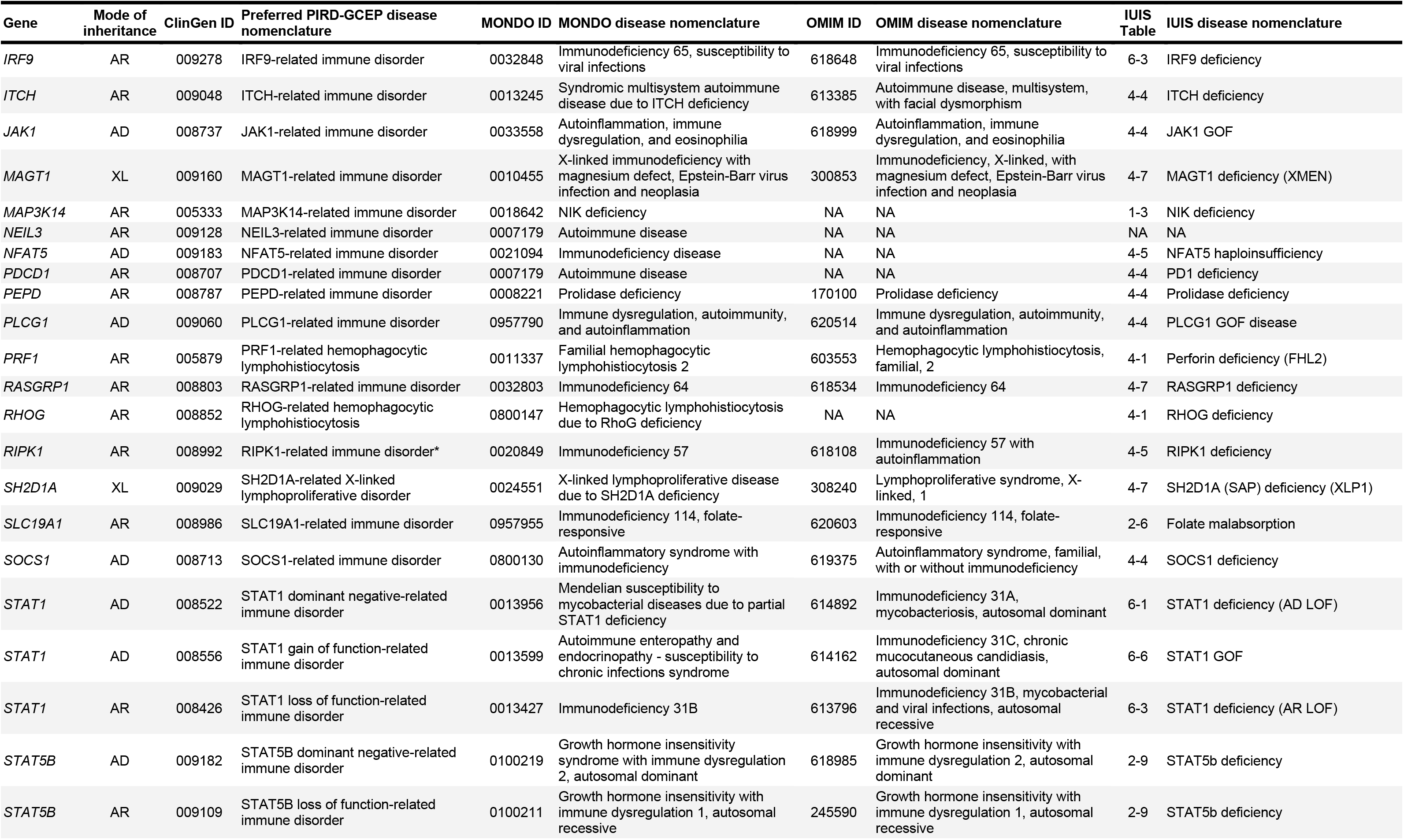
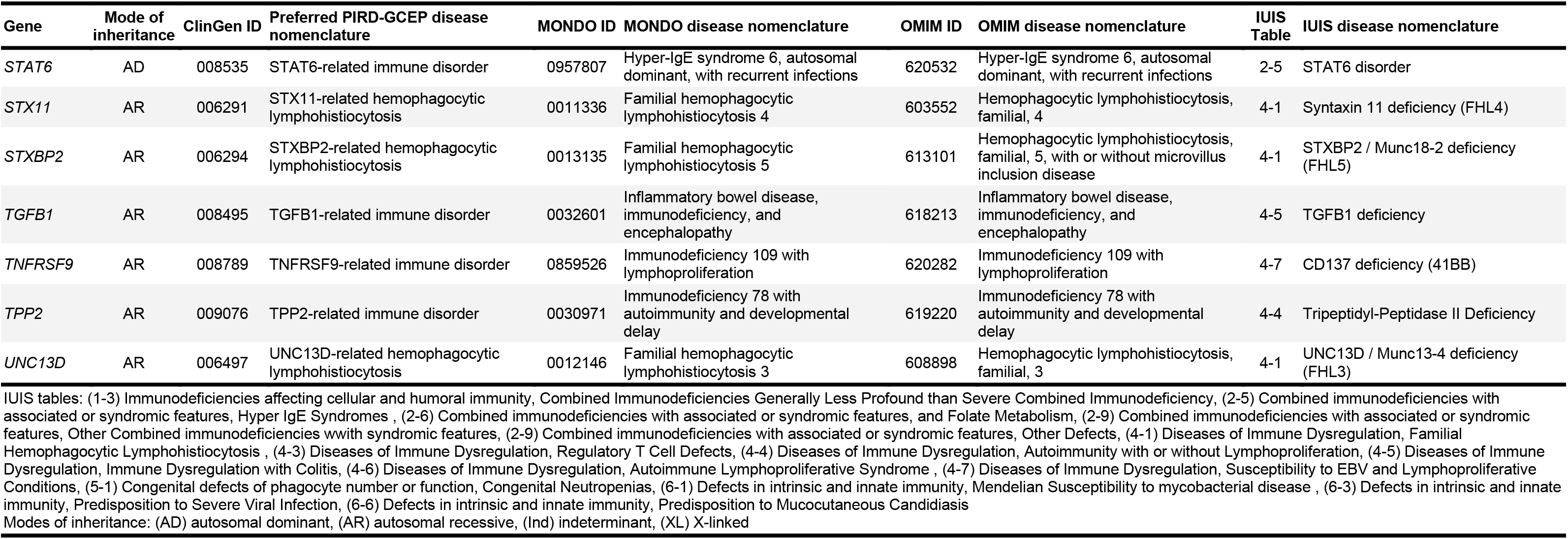
Unifying PIRD nomenclature.

